# Associations between Stress and Child Verbal Abuse and Corporal Punishment during the COVID-19 Pandemic and Potential Effect Modification by Lockdown Measures

**DOI:** 10.1101/2021.01.05.20248973

**Authors:** Rohani Jeharsae, Manusameen Jae-noh, Haneefah Jae-a-lee, Suhaida Waeteh, Nisuraida Nimu, Corliyoh Chewae, Malinee Yama, Nurin Dureh, Wit Wichaidit

**Affiliations:** Faculty of Nursing Pattani Campus, Prince of Songkla University, Mueang Pattani, Thailand; Pattani Provincial Public Health Office, Mueang Pattani, Thailand; Faculty of Science and Technology, Prince of Songkla University, Mueang Pattani, Thailand; Epidemiology Unit, Faculty of Medicine, Prince of Songkla University. Hat Yai, Thailand

**Keywords:** Child abuse, COVID-19, Lockdown, Stress, Hardship

## Abstract

**Background:** Child abuse appears to be on the increase during the COVID-19 pandemic, but the extent that lockdown measures modified the association between stress and abuses has not been systematically assessed.

**Objectives:** To assess: 1) the association between caregiver’s stress and self-reported verbal abuse and corporal punishment of a child in the household, and; 2) modification of the stated association by experienced COVID-19 lockdown measures.

**Participants and settings:** Caregivers residing in villages on lockdown in the Deep South of Thailand (n=466 participants)

**Methods:** We randomly sampled 12 villages in the study area, and 40 households per village. Trained enumerators who were residents of the sampled villages collected the data using phone-based interview. We measured stress level using the standard ST-5 questionnaire. We developed and pilot-tested questions for measurement of child abuse and lockdown experiences specifically for this study.

**Results:** Caregivers with moderate and higher levels of stress were more likely than caregivers with low level of stress to report verbal abuse (48% vs. 23%, respectively; Adj. OR = 3.12, 95% CI = 1.89, 5.15) and corporal punishment (28% vs. 8%, respectively; Adj. OR = 2.76, 95% CI = 1.41, 5.42). We found that COVID-19 lockdown experiences modified the associations between stress and verbal abuse and corporal punishment.

**Conclusion:** There were associations between stress and abuses, which were modified by lockdown experiences. However, social desirability, lack of details in the answers, and potential confounding by mental illness co-morbidities were notable limitations of the study. Caveat is advised in the interpretation of the study findings.

## INTRODUCTION

Child abuse and victimization of children appears to have increased during the COVID-19 pandemic (Fore & Cappa, 2020; Petrowski et al., 2020; Ramaswamy & Seshadri, 2020). Pandemic control (“lockdown”) measures commonly include school closure. As school staff commonly serve as screeners and reporters of child abuse and neglect, the occurrence of child abuses during lockdown is likely to be under-reported (Barboza et al., 2020; Caron et al., 2020). In the household, children can suffer verbal and physical abuses at the hands of their caregivers (Babvey et al., 2020; Barboza et al., 2020; Brown et al., 2020; Kovler et al., 2020; Lawson et al., 2020; Tener et al., 2020). COVID-19 lockdown can result in job loss (Lawson et al., 2020), loss of income, and material hardship (Xu et al., 2020), which are associated with parenting stress (Xu et al., 2020) and acts of child abuse (Barboza et al., 2020; Lawson et al., 2020).

There have been calls for suggestions of strategies to mitigate the effects of lockdown on child abuse (Fore & Cappa, 2020; Holmes et al., 2020; Kovler et al., 2020). Reframing coping mechanism, an individualized internal factor, has been shown to mitigate the effect of job loss on child abuse (Lawson et al., 2020). However, no study has directly measured the association between stress experienced by caregivers and self-reported acts of child abuse in low and middle-income country settings. Moreover, no study has assessed how COVID-19 lockdown measures, i.e., external factors present at the community and societal level, can modify the effect of stress on acts of child abuse.

Thailand is a middle-income country in South East Asia. Thailand has one of the biggest informal economies in the world, with approximately half of the economy in the informal sector (Buddhari & Rugpenthum, 2019). Restrictions of movement can immediately affect the population’s ability to earn livelihood. During the early phase of the pandemic, the Thai government declared a state of emergency in late March 2020 (Thai Ministry of Foreign Affairs, 2020). The government authorized provincial governors to implement lockdown at locations where COVID-19 cases are found. These measures included restriction of movement between communities, school closure, and restriction of business activities. However, governors were allowed to modify the implementation of these measures as appropriate for each specific location. Analyses of data from community-based survey of caregivers in these outbreak areas can provide potentially useful basic information for relevant stakeholders. The objectives of this study were to assess: 1) the extent that caregiver’s self-reported stress is associated with self-reported verbal abuse and corporal punishment of a child in the household, and; 2) the extent that this relationship varied according to experienced COVID-19 lockdown measures.

## METHODS

### Study Design and Setting

This cross-sectional study was part of a mixed-method rapid assessment survey of the situation of children during COVID-19 lockdown in the Deep South of Thailand (Jeharsae et al., 2020). The investigators collected data from villages on lockdown in the provinces of Pattani, Narathiwat and Yala, near Thailand’s border with Malaysia of approximately 7600 sq.km. in total size and total population of approximately 2 million.

### Study Instrument

The study instrument was a web-based interview questionnaire based on the Google Form platform. The questionnaire included 65 questions divided into 6 sections: 1) Information about the interviewer (for quality control purpose); 2) Characteristics of the participants; 3) Child care practices during the COVID-19 pandemic; 4) Impact of COVID-19 pandemic on the child; 5) Awareness of the impact of the COVID-19 pandemic on children, and; 6) Stress assessment. With the exception of the stress assessment section, all sections of the questionnaire were developed by the investigators. Three experts assessed the study instrument for content validity after its development. The investigators also pilot-tested the questionnaire online to assess reliability before actual data collection.

### Exposure Measurement: Self-Reported Stress

Self-reported stress was measured by interview in the Stress Assessment section of the questionnaire. The section was adapted from the Srithanya Stress Scale-5 Items (ST-5), a Thai language instrument designed for community-based self-assessment of emotional stress (Silpakit, 2008). Our slightly modified version of the ST-5 was adopted for interview with the following introduction and questions (“The following is a stress assessment questionnaire. Please answer as truthfully as possible. Think of how you have felt during the past 2 weeks. [How often did you…]: 1) Have problem sleeping, not being able to sleep or sleeping too much; 2) Have decreased concentration; 3) Feel irritable; 4) Feel bored; 5) Did not want to see anyone”). Each question had 4 possible choices (“almost none” (0 point), “sometimes” (1 point), “frequently” (2 points), “regularly” (3 points)).

We summed the points on each question to obtain the ST-5 score. As per official guidelines (Department of Mental Health, 2016), we considered participants who had 0-4 points on our modified ST-5 as having “Low” level of stress, participants with 5-7 points as having “Moderate” level of stress, participants with 8-9 points as having “High” level of stress, and participants with 10-15 points as having “Severe” level of stress.

### Outcome Measurement: Verbal Abuse of Children

We measured self-reported verbal abuse of child(ren) under 18 years of age by caregivers living in the same household by the first question in Section 4 (Impact of COVID-19 pandemic on the child), which was “During the past 1 month, have you used profanity or scolded a child aged under 18 years because you were stressed from economic conditions?”. The question had four possible answer choices: 1) “Never”; 2) “Yes, 1-3 times per week”; 3) “Yes, 4-6 times per week”, and; 4) “Yes, daily”. We considered participants who answered “Never” as those who did not self-report verbal abuse of a child in the household, and everyone else as those who did. We intended to treat those who did not provide an answer as having missing data, but all respondents answered the question.

### Outcome Measurement: Corporal Punishment of Children

In the second question of Section 4 (Impact of COVID-19 pandemic on the child), we asked “During the past 1 month, have you punished a child aged under 18 years by hitting because you were stressed from economic conditions?”. The question had the same four possible answer choices as questions on verbal abuse. We considered participants who answered “Never” as those who did not self-report corporal punishment of a child in the household, and everyone else as those who did so. We planned to treat those who did not provide an answer as having missing data, but all respondents answered the question.

### Effect Modifier: Experienced Lockdown Measures

In the study area, residents of a village that was “on lockdown” were not allowed to travel to another village except when purchasing provisions or seeing doctors. Residents were allowed to conduct trade within the village, but authorities would not allow any person to enter or leave the village premise.

We measured lockdown measures experienced by caregivers based on three questions in Section 2 (Characteristics of the participants): 1) “Is your village under quarantine or restriction of movement (no one moves into or out of the village)?”; 2) “Is someone from your household currently in quarantine at a quarantine site?”, and; 3) “Has someone in your household been sick with COVID-19?”. All questions had binary (“Yes” vs. “No”) responses. We divided participants into 3 “Lockdown Groups”: Lockdown Group 1 (those whose village was not under quarantine or restriction of movement); Lockdown Group 2 (those living in villages on COVID-19 quarantine or restriction of movement but no household member was quarantined or infected), and; Lockdown Group 3 (those living in villages on COVID-19 quarantine or restriction of movement and someone in the household was quarantined or infected). When we assessed the association between our exposure and outcomes, we stratified all analyses by lockdown groups to assess effect modification accordingly.

### Data Collection

We collected data during the month of June 2020, three months after the first lockdown was announced in Thailand. The investigators first obtained a list of 98 villages on lockdown in the study area from the provincial authorities’ announcements. The investigators then sampled 4 villages from each province using stratified random sampling. The investigators contacted the Sub-District Health Promotion Hospital serving the selected villages to obtain the list of households in the village, filtered only households with at least one child aged less than 18 years according to the Health Promotion Hospital’s database, and selected 40 of the filtered households using simple random sampling, thus we sampled a total of 480 households.

The investigators then asked Health Promotion Hospital staff to recruit field enumerators from residents of the targeted villages with at least a secondary school education who possessed their own smart phone knew how to access the internet and use a phone-based internet browser. The lead investigator then provided distance briefing for the enumerators, including the study protocols, research ethics, and social distancing protocols in case of need to conduct face-to-face interviews.

On the day of data collection. Health Promotion Hospital staff then provided the enumerators with the phone number of the heads of sampled households in their village. The enumerators then called the number and asked for the head of household’s informed consent and permission to conduct the survey. Heads of household gave informed consent or declined participation verbally by phone. Whence the head of household gave verbal informed consent, the enumerator then asked to speak to a family member who was deemed to be most closely involved in the care of child(ren) aged under 18 years in the household. If the sampled household did not have a listed phone number in their record, the enumerators were to visit the household while following social distancing protocol and other proscribed precautionary measures. However, such measure was not needed during data collection and all interviews were conducted via phone conversations.

### Statistical Analyses

We described the characteristics of the study participants, including prevalence of the exposure and outcomes, using descriptive statistics. We assessed the association between stress level and verbal abuse, and the association between stress level and corporal punishment, using univariate and multivariate logistic regression analyses, stratified by level of experienced hardship to assess effect modification. Given the unique characteristics of the social and economic contexts of the pandemic and the lack of previous findings from the study area, we decided to use a data-driven approach to identify confounders instead of using findings from studies conducted *a priori*. We assessed associations between participants’ basic characteristics and prevalence of the outcomes, and included all variables whose p-value of the association was less than 0.15 in the respective multivariate logistic regression models. Whereas inclusion of all variables resulted in malfunctions of the multivariate model, we excluded variables that crashed the model before obtaining the final multivariate model. For quality control purpose, we decided to exclude participants who indicated that their sub-district did not have a COVID-19 infection from all analyses.

## RESULTS

Among the 480 sampled households, 473 head of households gave verbal consent and caregivers in the household agreed to participate. Among the 473 caregiver participants, there were 7 participants who indicated that their sub-district did not have someone infected with COVID-9 and were excluded from the study (n=466 participants). The majority of the participants were women and a parent of at least one child in the household. Participants were mostly Muslims, married, had secondary education or less, had household monthly income of 10,000 THB or less, and the primary source of the income was not from fixed salary (*Table 1*). The majority of participants lived in a village under lockdown, although a small minority reported that someone in their household was quarantined or infected with COVID-19. Most participants reported low level of stress during the past month. Slightly more than one-fourth of participants reported verbal abuse of a child in the past month, and one-tenth reported corporal punishment.

**Table 1.**
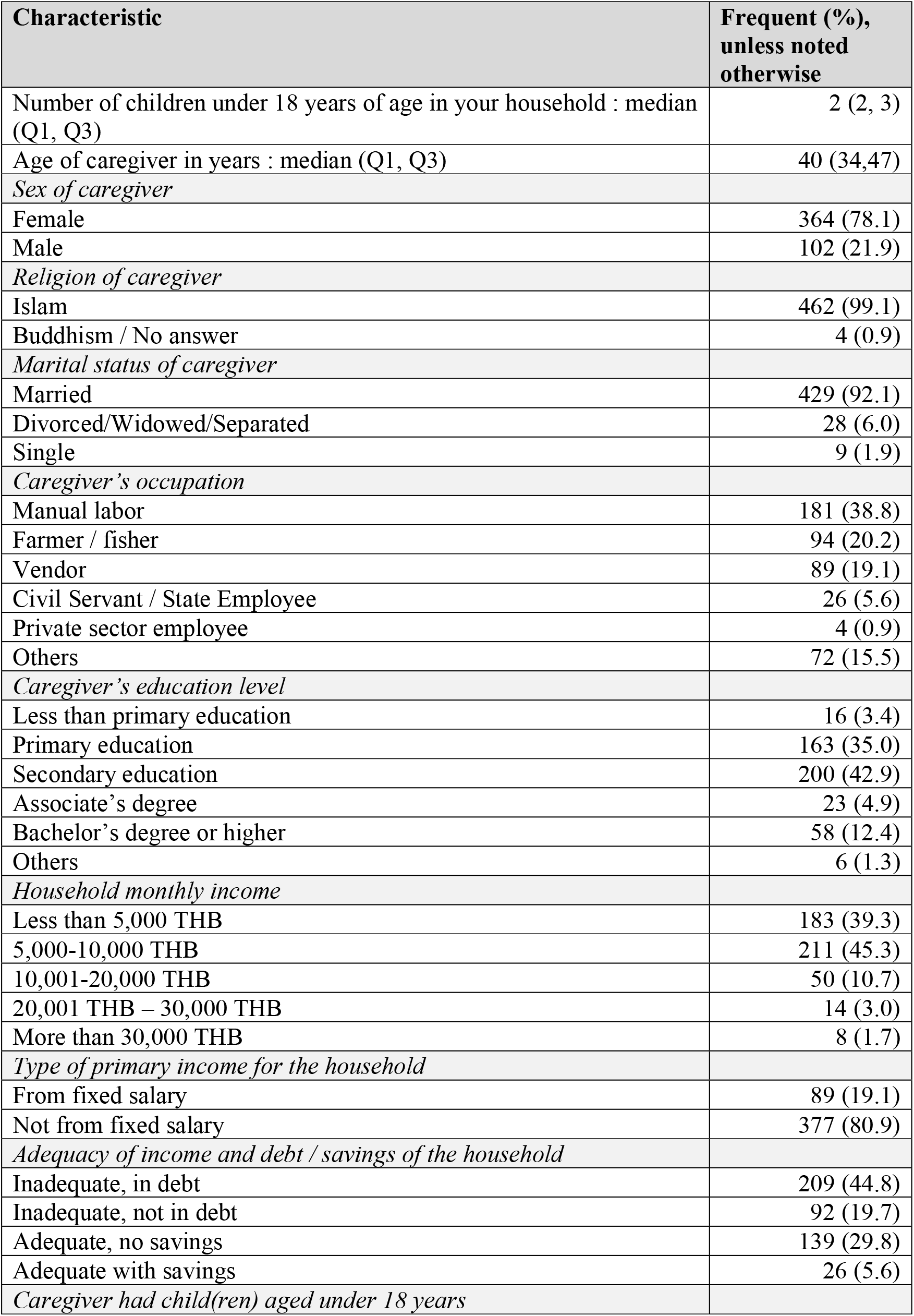
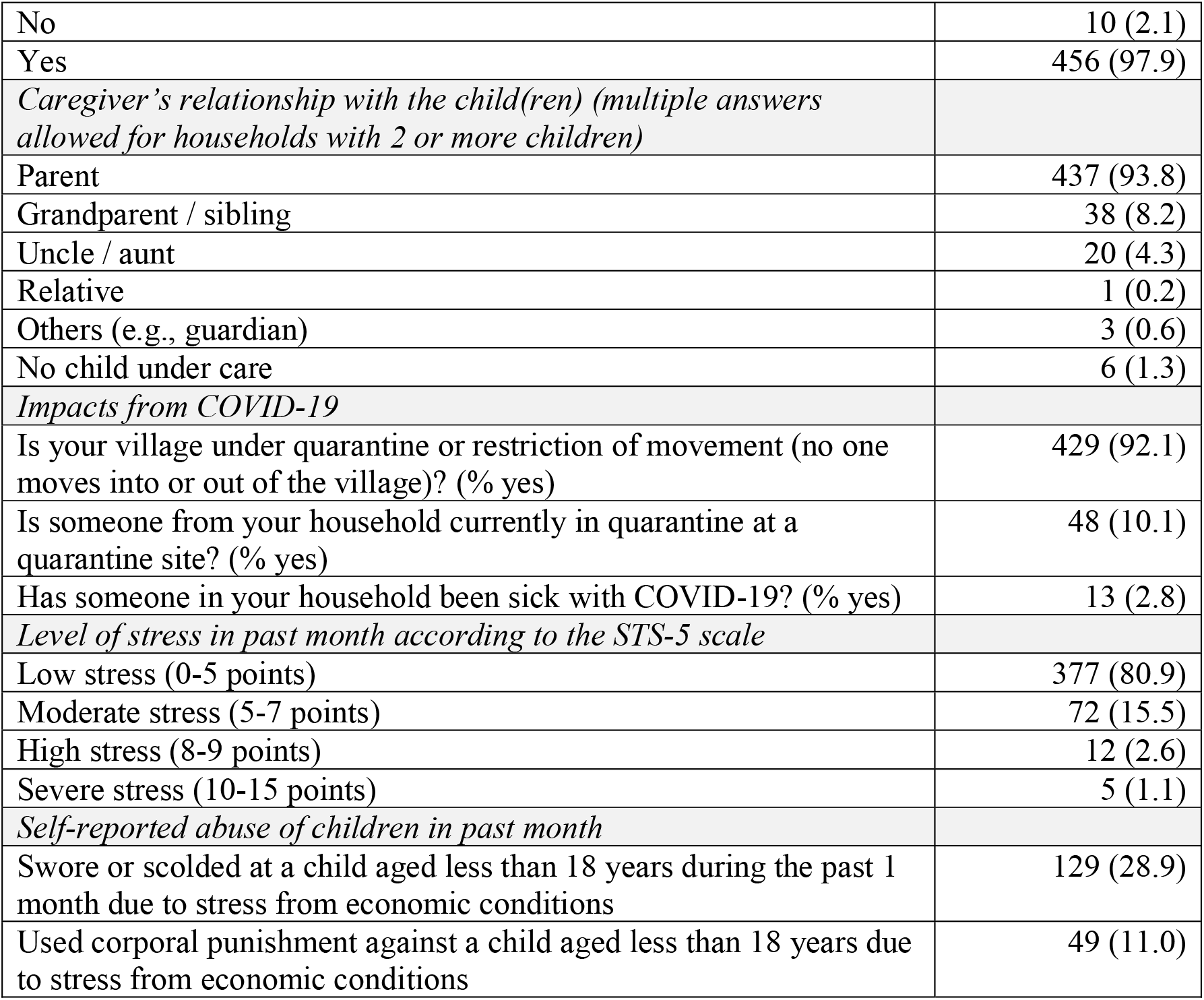
Basic Characteristics of the Study Participants and the Participants’ Household, Stratified by Stress Level (n= 466 caregivers)

| **Characteristic** | **Frequent (%), unless noted otherwise** |
| --- | --- |
| Number of children under 18 years of age in your household : median (Q1, Q3) | 2 (2, 3) |
| Age of caregiver in years : median (Q1, Q3) | 40 (34,47) |
| *Sex of caregiver* |  |
| Female | 364 (78.1) |
| Male | 102 (21.9) |
| *Religion of caregiver* |  |
| Islam | 462 (99.1) |
| Buddhism / No answer | 4 (0.9) |
| *Marital status of caregiver* |  |
| Married | 429 (92.1) |
| Divorced/Widowed/Separated | 28 (6.0) |
| Single | 9 (1.9) |
| *Caregiver's occupation* |  |
| Manual labor | 181 (38.8) |
| Farmer / fisher | 94 (20.2) |
| Vendor | 89 (19.1) |
| Civil Servant / State Employee | 26 (5.6) |
| Private sector employee | 4 (0.9) |
| Others | 72 (15.5) |
| *Caregiver's education level* |  |
| Less than primary education | 16 (3.4) |
| Primary education | 163 (35.0) |
| Secondary education | 200 (42.9) |
| Associate's degree | 23 (4.9) |
| Bachelor's degree or higher | 58 (12.4) |
| Others | 6 (1.3) |
| *Household monthly income* |  |
| Less than 5,000 THB | 183 (39.3) |
| 5,000-10,000 THB | 211 (45.3) |
| 10,001-20,000 THB | 50 (10.7) |
| 20,001 THB – 30,000 THB | 14 (3.0) |
| More than 30,000 THB | 8 (1.7) |
| *Type of primary income for the household* |  |
| From fixed salary | 89 (19.1) |
| Not from fixed salary | 377 (80.9) |
| *Adequacy of income and debt / savings of the household* |  |
| Inadequate, in debt | 209 (44.8) |
| Inadequate, not in debt | 92 (19.7) |
| Adequate, no savings | 139 (29.8) |
| Adequate with savings | 26 (5.6) |
| *Caregiver had child(ren) aged under 18 years* |  |
| No | 10 (2.1) |
| Yes | 456 (97.9) |
| *Caregiver's relationship with the child(ren) (multiple answers allowed for households with 2 or more children)* |  |
| Parent | 437 (93.8) |
| Grandparent / sibling | 38 (8.2) |
| Uncle / aunt | 20 (4.3) |
| Relative | 1 (0.2) |
| Others (e.g., guardian) | 3 (0.6) |
| No child under care | 6 (1.3) |
| *Impacts from COVID-19* |  |
| Is your village under quarantine or restriction of movement (no one moves into or out of the village)? (% yes) | 429 (92.1) |
| Is someone from your household currently in quarantine at a quarantine site? (% yes) | 48 (10.1) |
| Has someone in your household been sick with COVID-19? (% yes) | 13 (2.8) |
| *Level of stress in past month according to the STS-5 scale* |  |
| Low stress (0-5 points) | 377 (80.9) |
| Moderate stress (5-7 points) | 72 (15.5) |
| High stress (8-9 points) | 12 (2.6) |
| Severe stress (10-15 points) | 5 (1.1) |
| *Self-reported abuse of children in past month* |  |
| Swore or scolded at a child aged less than 18 years during the past 1 month due to stress from economic conditions | 129 (28.9) |
| Used corporal punishment against a child aged less than 18 years due to stress from economic conditions | 49 (11.0) |

Self-reported verbal abuse of children was positively associated with number of children in the household, household primary income not coming from fixed salary, and perceived inadequacy of household income (*Table 2*). Verbal abuse was negatively associated with caregiver’s higher level of education. The same patterns of association existed with regard to self-reported corporal punishment of children (*Table 3*).

**Table 2.** Characteristics of participants associated with self-reported verbal abuse of children within past month among primary caregivers (n= 466 Caregivers)

| Characteristics | No verbal abuse (n=337)<br>n (%) | Verbal abuse (n=129)<br>n (%) | P-value |
| --- | --- | --- | --- |
| Number of children under 18 years of age in your household (median (Q1,Q3)) | 2 (1, 3) | 2 (2, 3) | **0.014** |
| Age of caregiver (years) (median (Q1,Q3)) | 41 (34, 47) | 40 (33, 46) | 0.235 |
| *Sex of caregiver* |  |  |  |
| Female (n=364) | 258 (70.9) | 106 (29.1) | 0.236 |
| Male (n=102) | 79 (77.5) | 23 (22.5) |  |
| *Religion of caregiver* |  |  |  |
| Islam (n=462) | 335 (72.5) | 127 (27.5) | 0.307 |
| Buddhism / No answer (n=4) | 2 (50.0) | 2 (50.0) |  |
| *Marital status of caregiver* |  |  |  |
| Married (n=429) | 308 (71.8) | 121 (28.2) | 0.173 |
| Divorced/Widowed/Separated (n=28) | 20 (71.4) | 8 (28.6) |  |
| Single (n=9) | 9 (100.0) | 0 (0.0) |  |
| *Caregiver's occupation* |  |  |  |
| Manual labor (n=181) | 131 (72.4) | 50 (27.6) | 0.879 |
| Farmer / fisher (n=94) | 69 (73.4) | 25 (26.6) |  |
| Vendor (n=89) | 63 (70.8) | 26 (29.2) |  |
| Civil Servant / State Employee (n=26) | 19 (73.1) | 7 (26.9) |  |
| Private sector employee (n=4) | 4 (100.0) | 0 (0.0) |  |
| Others (n=72) | 51 (70.8) | 21 (29.2) |  |
| *Caregiver's education level* |  |  |  |
| Less than primary education (n=16) | 9 (56.2) | 7 (43.8) | **0.040** |
| Primary education (n=163) | 125 (76.7) | 38 (23.3) |  |
| Secondary education (n=200) | 132 (66.0) | 68 (34.0) |  |
| Associate's degree (n=23) | 18 (78.3) | 5 (21.7) |  |
| Bachelor's degree or higher (n=58) | 48 (82.8) | 10 (17.2) |  |
| Others (n=6) | 5 (83.3) | 1 (16.7) |  |
| *Household monthly income* |  |  |  |
| Less than 5,000 THB (n=183) | 128 (69.9) | 55 (30.1) | 0.906 |
| 5,000-10,000 THB (n=211) | 155 (73.5) | 56 (26.5) |  |
| 10,001-20,000 THB (n=50) | 37 (74.0) | 13 (26.0) |  |
| 20,001 THB – 30,000 THB (n=14) | 11 (78.6) | 3 (21.4) |  |
| More than 30,000 THB (n=8) | 6 (75.0) | 2 (25.0) |  |
| *Type of primary income for the household* |  |  |  |
| From fixed salary (n=89) | 73 (82.0) | 16 (18.0) | **0.032** |
| Not from fixed salary (n=377) | 264 (70.0) | 113 (30.0) |  |
| *Perceived adequacy of income / household financial situation* |  |  |  |
| Inadequate, in debt (n=209) | 150 (71.8) | 59 (28.2) | **0.136** |
| Inadequate, not in debt (n=92) | 59 (64.1) | 33 (35.2) |  |
| Adequate, no savings (n=139) | 107 (77.0) | 32 (23.0) |  |
| Adequate with savings (n=26) | 21 (80.8) | 5 (19.2) |  |

| *Caregiver had child(ren) aged under 18 years* |  |  |  |
| --- | --- | --- | --- |
| No (n=10) | 6 (60.0) | 4 (40.0) | 0.601 |
| Yes (n=456) | 331 (72.6) | 125 (27.4) |  |

Bold texts denote statistical significance at 95% level of confidence

**Table 3.**
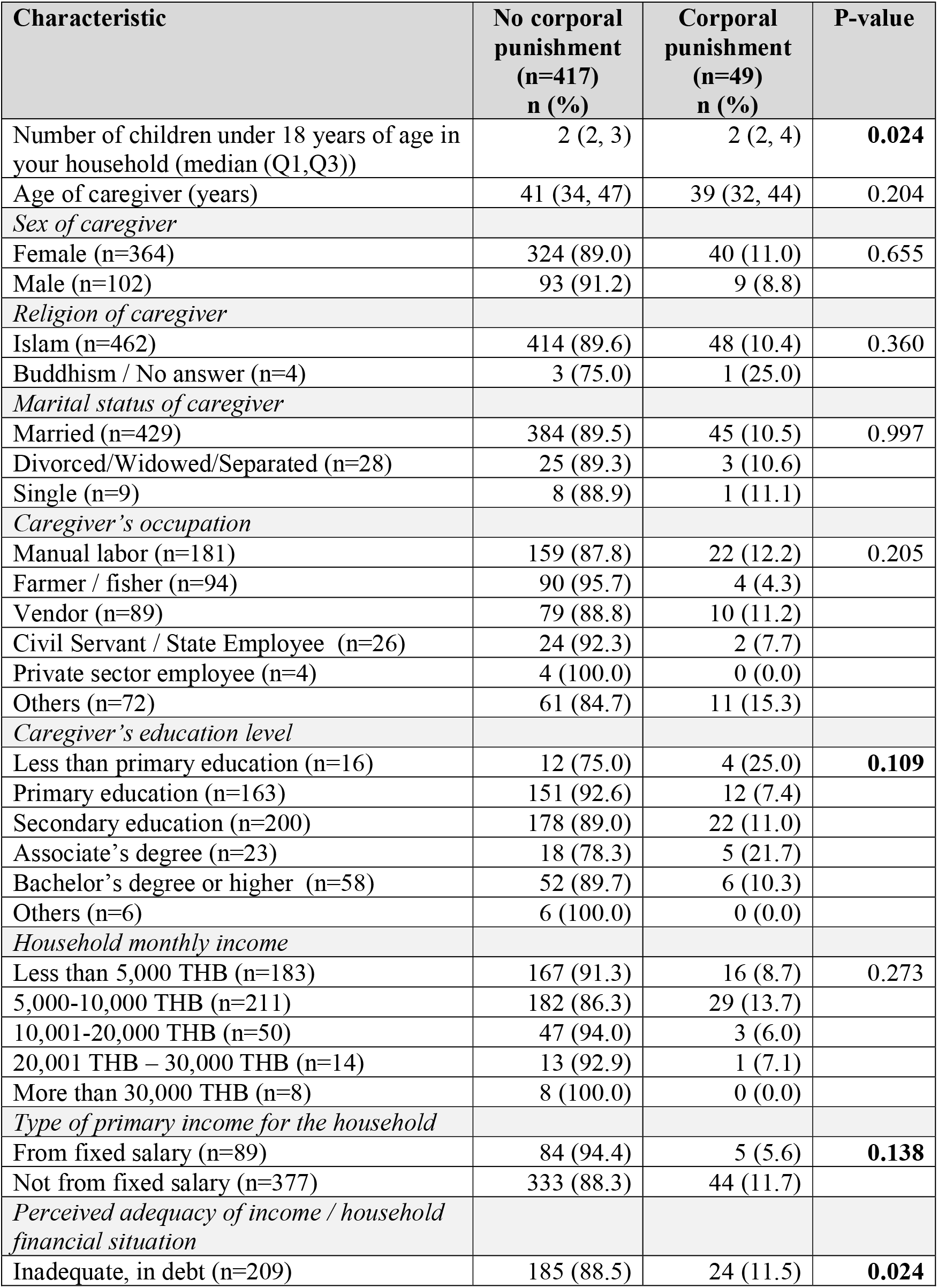
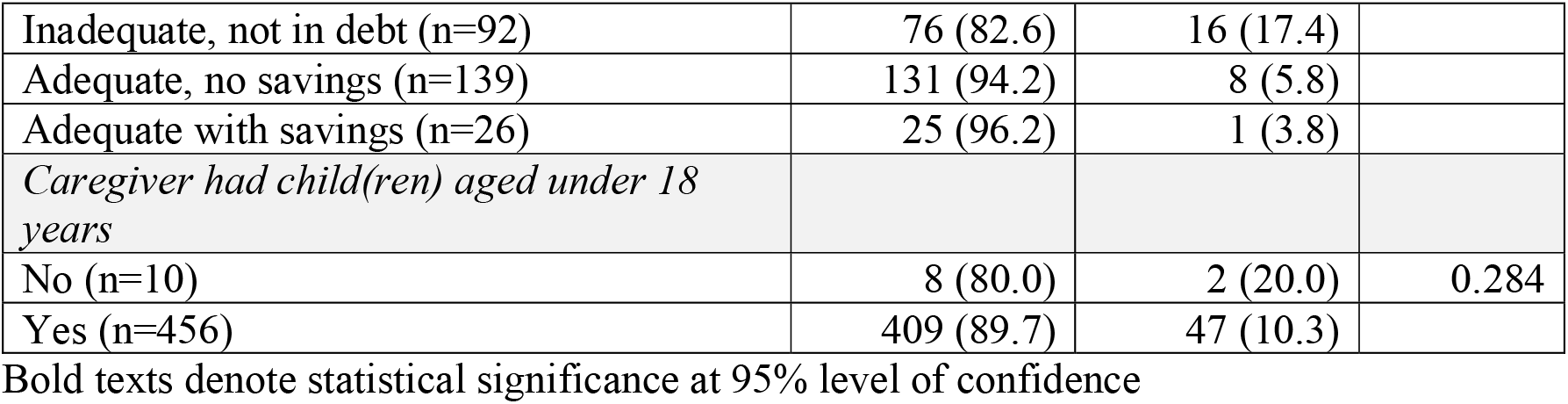
Characteristics of participants associated with self-reported corporal punishment of children among primary caregivers (n= 466 Caregivers)

Caregivers with moderate or higher level of stress during past month had 3.1 times higher odds of self-reported verbal abuse of children due to stress from economic conditions (*Table 4*). The association was strongest among caregivers in Lockdown Group 2 (living in villages on quarantine or restriction of movement but no household member was quarantined or infected) and weakest among caregivers in Lockdown Group 3 (living in villages on quarantine or restriction of movement and someone in the household was quarantined or infected). Similar patterns existed for corporal punishment of children due to stress from economic conditions (*Table 5*). Caregivers with moderate or higher level of stress during the past month had 2.8 times higher odds of corporal punishment. The association was strongest among caregivers in Lockdown Group 2 and weakest among caregivers in Lockdown Group 3.

**Table 4.** Association between stress level and self-reported verbal abuse of children among primary caregivers, stratified by experience (groups) of COVID-19 lockdown (n= 466 Caregivers)

| Stress Level | No verbal abuse<br>(n=337)<br>n (%) | Verbal abuse<br>(n=129)<br>n (%) | Crude OR<br>(95% CI) | Adjusted OR<br>(95% CI) |
| --- | --- | --- | --- | --- |
| *All caregivers (n=466)* |  |  |  |  |
| Low stress (Ref.) (n=377) | 291 (77.2) | 86 (22.8) | 1.0 (Ref.) | 1.0 (Ref.) |
| Moderate and higher stress<br>(n=89) | 46 (51.7) | 43 (48.3) | **3.16**<br>**(1.96,5.11)** | **3.12**<br>**(1.89,5.15)<sup>a</sup>** |
| *Caregivers in Lockdown Group 1 (n=37)<sup>d</sup>* |  |  |  |  |
| Low stress (Ref.) (n=32) | 22 (68.8%) | 10 (31.2%) | 1.0 (Ref.) | 1.0 (Ref.) |
| Moderate and higher stress<br>(n=5) | 2 (40.0%) | 3 (60.0%) | 3.30<br>(0.47,22.94) | 3.44<br>(0.41,29.1)<sup>b</sup> |
| *Caregivers in Lockdown Group 2 (n=379)<sup>d</sup>* |  |  |  |  |
| Low stress (Ref.) (n=304) | 243 (79.9%) | 61 (20.1%) | 1.0 (Ref.) | 1.0 (Ref.) |
| Moderate and higher stress<br>(n=75) | 39 (52.0%) | 36 (48.0%) | **3.68**<br>**(2.16,6.27)** | **3.52 (2,6.19)<sup>a</sup>** |
| *Caregivers in Lockdown Group 3 (n=50)<sup>d</sup>* |  |  |  |  |
| Low stress (Ref.) (n=41) | 26 (63.4%) | 15 (36.6%) | 1.0 (Ref.) | 1.0 (Ref.) |
| Moderate and higher stress<br>(n=9) | 5 (55.6%) | 4 (44.4%) | 1.39<br>(0.32,5.97) | 1.56<br>(0.31,7.96)<sup>c</sup> |

Bold texts denote statistical significance at 95 level of confidence
a. Adjusted for number of children in household, caregiver's education level, having household primary income from fixed salary, and perceived adequacy of income / household financial situation
b. Adjusted for number of children in household and perceived adequacy of income / household financial situation
c. Adjusted for number of children in household, having household primary income from fixed salary, and perceived adequacy of income / household financial situation
d. Lockdown Group 1 = living in villages not on COVID-19 quarantine or restriction of movement; Lockdown Group 2 = living in villages on COVID-19 quarantine or restriction of movement but no household member was quarantined or infected; Lockdown Group 3 = living in villages on COVID-19 quarantine or restriction of movement and someone in the household was quarantined or infected

**Table 5.** Association between stress level and self-reported corporal punishment of children among primary caregivers, stratified by experience (groups) of COVID-19 lockdown (n= 466 Caregivers)

| Stress Level | No corporal punishment (n=417)<br>n (%) | Corporal punishment (n=49)<br>n (%) | Crude OR (95% CI) | Adjusted OR (95% CI) |
| --- | --- | --- | --- | --- |
| *All caregivers (n=466)* |  |  |  |  |
| Low stress (Ref.) (n=377) | 346 (91.8) | 31 (8.2) | 1.0 (Ref.) | 1.0 (Ref.) |
| Moderate and higher stress (n=89) | 71 (79.8) | 18 (20.2) | **2.83**<br>**(1.5,5.34)** | **2.76**<br>**(1.41,5.42)<sup>a</sup>** |
| *Caregivers in Lockdown Group 1 (n=37)<sup>c</sup>* |  |  |  |  |
| Low stress (Ref.) (n=32) | 28 (87.5%) | 4 (12.5%) | 1.0 (Ref.) | 1.0 (Ref.) |
| Moderate and higher stress (n=5) | 4 (80.0%) | 1 (20.0%) | 1.75<br>(0.15,19.86) | 1.78<br>(0.15,21.34)<sup>b</sup> |
| *Caregivers in Lockdown Group 2 (n=379)<sup>c</sup>* |  |  |  |  |
| Low stress (Ref.) (n=304) | 282 (92.8%) | 22 (7.2%) | 1.0 (Ref.) | 1.0 (Ref.) |
| Moderate and higher stress (n=75) | 59 (78.7%) | 16 (21.3%) | **3.48**<br>**(1.72,7.02)** | **3.40**<br>**(1.6,7.21)<sup>a</sup>** |
| *Caregivers in Lockdown Group 3 (n=50)<sup>c</sup>* |  |  |  |  |
| Low stress (Ref.) (n=41) | 36 (87.8%) | 5 (12.2%) | 1.0 (Ref.) | 1.0 (Ref.) |
| Moderate and higher stress (n=9) | 8 (88.9%) | 1 (11.1%) | 0.90<br>(0.09,8.80) | 0.89<br>(0.09,8.81)<sup>b</sup> |

Bold texts denote statistical significance at 95% level of confidence
a. Adjusted for number of children in household, caregiver's education level, having household primary income from fixed salary, and perceived adequacy of income / household financial situation
b. Adjusted for number of children in household
c. Lockdown Group 1 = living in villages not on COVID-19 quarantine or restriction of movement; Lockdown Group 2 = living in villages on COVID-19 quarantine or restriction of movement but no household member was quarantined or infected; Lockdown Group 3 = living in villages on COVID-19 quarantine or restriction of movement and someone in the household was quarantined or infected

## DISCUSSION

We found that associations between caregivers’ stress level and verbal abuse and corporal punishment of a child in the household. We also found that experiencing lesser hardship (i.e., living in villages not on COVID-19 quarantine or restriction of movement, or living in villages on COVID-19 quarantine or restriction of movement but with quarantined or infected family members) buffered the stated associations. The findings of this study can potentially contribute to the understanding of child abuse in the context of COVID-19 pandemic, although several considerations should be made in the interpretation of the findings.

The Srithanya stress scale 5-items version (ST-5) was developed for rapid screening of stress in community settings in Thailand, which suited the context of our study. However, responses to the ST-5 correlates with those of tools used to measure depression and anxiety (Silpakit, 2008), thus those with higher score on the tool could have other mental health comorbidities that were not measured. Stress is correlated with anxiety and depression (Brown et al., 2020), and parents with anxiety and depression are more likely to maltreat their children (Brown et al., 2020; Lawson et al., 2020), thus it is possible that the effect of confounding by comorbidities could have influenced the observed findings. Furthermore, the answer choices in the adapted ST-5 tool regarding frequency of stress symptoms were non-specific and subjective to the respondent’s own perception. The perceived frequency could be under-estimated or over-estimated, which then affected the ST-5 score and stress categorization. If the inaccuracies were systematically different between outcome and no-outcome groups, then information bias could have been present in the findings. Future studies should consider providing concise definitions for the categories (e.g., “The term ‘Sometimes’ refers to between 3 and 7 occasions during the past 2 weeks”).

Our participants reported verbal abuse and corporal punishment due to stress, including among those who reported infrequent or no symptom of stress according to the ST-5. Such findings could have been influenced by differing windows of measurement for the outcomes and exposure (history in past month for verbal abuse or corporal punishment, and history in past two weeks for stress). In multiple settings, reports of child abuse decreased after lockdown implementation (Barboza et al., 2020; Caron et al., 2020; Martins-Filho et al., 2020; Rapoport et al., 2020) possibly due to school closure and consequent lack of reporting by school staff (Barboza et al., 2020; Caron et al., 2020; Fore & Cappa, 2020; Jacob, 2020; Rapoport et al., 2020; Thomas et al., 2020), despite worsening family dynamics(Tener et al., 2020) and drastic increase in contact with helplines(Petrowski et al., 2020). In our study, social desirability as well as the respondents’ own perception of whether stress was the attributing factor for the act of verbal abuse or corporal punishment could have led to under-reporting of the outcomes. In that regard, we decided to use the context of corporal punishment of children under 18 years in the household due to economic stress as proxy for direct question about physical abuse with the intention to circumvent the influence of social desirability. Although corporal punishments with extensive injury to the child can be considered physical abuse if the punishment results in extensive injury(Coleman et al., 2010), our measurement question did not include details of the punishment. Some acts of corporal punishment reported in our study might not be acts of child abuse, and caveat is advised in the interpretation of these findings.

The associations between stress and verbal abuse and corporal punishment were weakest among caregivers living in villages on quarantine or restriction of movement with quarantined or ill family members. In other words, there seem to be effect modification by lockdown experience, although we are not quite sure of the mechanism behind it. It is possible that some of the household’s children were quarantined or treated and placed in an area designated by the state with chaprones instead of staying at home, which reduced the amount of contact between the caregivers and the children and, subsequently, the opportunities where abuses could occur. It is also possible that lockdown experiences were associated with economic or material hardship, which was found to be associated with parenting stress (Xu et al., 2020). In that regard, we did not directly ask participants about the extent that they experienced material hardship, and future studies should consider making such measurements using tools recently developed for household surveys (Fallon et al., 2020). In addition, the study area has experienced by decades-long armed conflict with as many as 20,000 casualties(Deep South Watch, 2020). Casualty was particularly heavy during the first decade of the conflict (Abuza, 2015; Human Rights Watch, 2007). The security situation deterred investment and created a uniquely complex setting for this survey. Interpretation and generalization of findings from this study should consider these contexts accordingly.

This is one of the first studies to assess how lockdown measures modifies the association between stress and child abuses in a post-conflict setting. However, a number of limitations should be considered in the interpretation of the study findings. Firstly, the cross-sectional design of the study as well as the inconsistent windows of measurement for the exposure and outcomes precludes the ability to make causal inferences. Secondly, abuses tended to be under-reported, particularly by the abuser, thus the study findings are prone to social desirability bias. Prevalence of the outcomes also could have been affected by the subjectivity of what the respondents considered as acts of verbal abuse and corporal punishment attributable to stress. Lastly, mental illness co-morbidities could have accounted for parts or all of the observed associations, but these co-morbidities were not assessed. In other words, uncontrolled confounding could have accounted, at least partially, for the study findings.

## CONCLUSION

In a phone-based survey of residents in sub-districts of southern Thailand with COVID-19 cases, we found association between level of stress among caregivers and self-reported verbal abuse and corporal punishment of children in their households, and that living in villages without quarantine or restriction of movement and having family members under quarantine or infected with COVID-19 mitigated the strength of these associations. However, a number of limitations existed in the study, including potential under-reporting due to social desirability, lack of details and ability to draw causal inferences, and potential confounding by mental illness co-morbidities. Caveat is advised in the interpretation of the study findings.

## Data Availability

Data Availability Statement: Study data available from the corresponding author (WW) upon reasonable request.

## Acknowledgement

We would like to thank all data collectors to worked tirelessly amidst a setting of armed conflict combined with the COVID-19 pandemic. We also would like to thank Ms. Jirawan Jayuphan for her statistical consultation throughout the data analysis process and UNICEF Thailand Country Office for providing financial support

## REFERENCES

Abuza, Z. (2015). The Smoldering Thai Insurgency. CTC Sentinel, 8(6), 8–11.

Babvey, P., Capela, F., Cappa, C., Lipizzi, C., Petrowski, N., & Ramirez-Marquez, J. (2020). Using social media data for assessing children’s exposure to violence during the COVID-19 pandemic. Child Abuse & Neglect, 104747. PMC. https://doi.org/10.1016/j.chiabu.2020.104747

Barboza, G. E., Schiamberg, L. B., & Pachl, L. (2020). A spatiotemporal analysis of the impact of COVID-19 on child abuse and neglect in the city of Los Angeles, California. Child Abuse & Neglect, 104740–104740. PubMed. https://doi.org/10.1016/j.chiabu.2020.104740

Brown, S. M., Doom, J. R., Lechuga-Peña, S., Watamura, S. E., & Koppels, T. (2020). Stress and parenting during the global COVID-19 pandemic. Child Abuse & Neglect, 104699–104699. PubMed. https://doi.org/10.1016/j.chiabu.2020.104699

Buddhari, A., & Rugpenthum, P. (2019). FAQ Focused and Quick: A Better Understanding of Thailand’s Informal Sector. Bank of Thailand. https://www.bot.or.th/Thai/MonetaryPolicy/ArticleAndResearch/FAQ/FAQ_156.pdf

Caron, F., Plancq, M.-C., Tourneux, P., Gouron, R., & Klein, C. (2020). Was child abuse underdetected during the COVID-19 lockdown? Archives de Pediatrie?: Organe Officiel de La Societe Francaise de Pediatrie, 27(7), 399–400. PubMed. https://doi.org/10.1016/j.arcped.2020.07.010

Coleman, D. L., Dodge, K. A., & Campbell, S. K. (2010). WHERE AND HOW TO DRAW THE LINE BETWEEN REASONABLE CORPORAL PUNISHMENT AND ABUSE. Law and Contemporary Problems, 73(2), 107–166. PubMed.

Deep South Watch. (2020). Summary of Incidents in Southern Thailand, FEBRUARY 2020. https://deepsouthwatch.org/sites/default/files/blogs/attachment/datasheet-022020-en.pdf

Department of Mental Health. (2016). Stress Assessment Questionnaire (ST5). Department of Mental Health, Thai Ministry of Public Health. https://www.dmh.go.th/test/qtest5/

Fallon, B., Lefebvre, R., Collin-Vézina, D., Houston, E., Joh-Carnella, N., Malti, T., Filippelli, J., Schumaker, K., Manel, W., Kartusch, M., & Cash, S. (2020). Screening for economic hardship for child welfare-involved families during the COVID-19 pandemic: A rapid partnership response. Child Abuse & Neglect, 104706–104706. PubMed. https://doi.org/10.1016/j.chiabu.2020.104706

Fore, H. H., & Cappa, C. (2020). Violence against Children in the Time of COVID-19: What we have learned, what remains unknown and the opportunities that lie ahead. Child Abuse & Neglect, 104776. PMC. https://doi.org/10.1016/j.chiabu.2020.104776

Holmes, E. A., O’Connor, R. C., Perry, V. H., Tracey, I., Wessely, S., Arseneault, L., Ballard, C., Christensen, H., Cohen Silver, R., Everall, I., Ford, T., John, A., Kabir, T., King, K., Madan, I., Michie, S., Przybylski, A. K., Shafran, R., Sweeney, A., … Bullmore, E. (2020). Multidisciplinary research priorities for the COVID-19 pandemic: A call for action for mental health science. The Lancet. Psychiatry, 7(6), 547–560. PubMed. https://doi.org/10.1016/S2215-0366(20)30168-1

Human Rights Watch. (2007). No One Is Safe: Insurgent Attacks on Civilians in Thailand’s Southern Border Provinces: II. A Brief History of Insurgency in the Southern Border Provinces. http://www.hrw.org/reports/2007/thailand0807/3.htm

Jacob, H. (2020). Safeguarding Children in a Pandemic: Pandemonium with Possibility? Child Abuse Review (Chichester, England?: 1992), 10.1002/car.2654. PubMed. https://doi.org/10.1002/car.2654

Jeharsae, R., Jaenoh, M., Jah-a-lee, H., Waeteh, S., Dureh, N., Nimu, N., Yama, M., & Chewae, K. (2020). Rapid Assessment of Impacts of COVID-19 Pandemic on Children in Lockdown Villages in Deep South of Thailand (p. 38). UNICEF and Prince of Songkla University.

Kovler, M. L., Ziegfeld, S., Ryan, L. M., Goldstein, M. A., Gardner, R., Garcia, A. V., & Nasr, I. W. (2020). Increased proportion of physical child abuse injuries at a level I pediatric trauma center during the Covid-19 pandemic. Child Abuse & Neglect, 104756–104756. PubMed. https://doi.org/10.1016/j.chiabu.2020.104756

Lawson, M., Piel, M. H., & Simon, M. (2020). Child Maltreatment during the COVID-19 Pandemic: Consequences of Parental Job Loss on Psychological and Physical Abuse Towards Children. Child Abuse & Neglect, 104709–104709. PubMed. https://doi.org/10.1016/j.chiabu.2020.104709

Martins-Filho, P. R., Damascena, N. P., Lage, R. C., & Sposato, K. B. (2020). Decrease in child abuse notifications during COVID-19 outbreak: A reason for worry or celebration? Journal of Paediatrics and Child Health, 10.1111/jpc.15213. PubMed. https://doi.org/10.1111/jpc.15213

Petrowski, N., Cappa, C., Pereira, A., Mason, H., & Daban, R. A. (2020). Violence against children during COVID-19 Assessing and understanding change in use of helplines. Child Abuse & Neglect, 104757. PMC. https://doi.org/10.1016/j.chiabu.2020.104757

Ramaswamy, S., & Seshadri, S. (2020). Children on the brink: Risks for child protection, sexual abuse, and related mental health problems in the COVID-19 pandemic. Indian Journal of Psychiatry, 62(Suppl 3), S404–S413. PubMed. https://doi.org/10.4103/psychiatry.IndianJPsychiatry_1032_20

Rapoport, E., Reisert, H., Schoeman, E., & Adesman, A. (2020). Reporting of child maltreatment during the SARS-CoV-2 pandemic in New York City from March to May 2020. Child Abuse & Neglect, 104719–104719. PubMed. https://doi.org/10.1016/j.chiabu.2020.104719

Silpakit, O. (2008). Srithanya stress scale. Journal of Mental Health of Thailand, 16(3), 177– 185.

Tener, D., Marmor, A., Katz, C., Newman, A., Silovsky, J. F., Shields, J., & Taylor, E. (2020). How does COVID-19 impact intrafamilial child sexual abuse? Comparison analysis of reports by practitioners in Israel and the US. Child Abuse & Neglect, 104779–104779. PubMed. https://doi.org/10.1016/j.chiabu.2020.104779

Thai Ministry of Foreign Affairs. (2020). Official Statement of the Office of the Prime Minister RE?: Declaration of an Emergency Situation pursuant to the Emergency Decree on Public Administration in Emergency Situations B.E. 2548 (2005). http://www.mfa.go.th/main/contents/files/news3-20200326-211539-804409.pdf

Thomas, E. Y., Anurudran, A., Robb, K., & Burke, T. F. (2020). Spotlight on child abuse and neglect response in the time of COVID-19. The Lancet. Public Health, 5(7), e371– e371. PubMed. https://doi.org/10.1016/S2468-2667(20)30143-2

Xu, Y., Wu, Q., Levkoff, S. E., & Jedwab, M. (2020). Material hardship and parenting stress among grandparent kinship providers during the COVID-19 pandemic: The mediating role of grandparents’ mental health. Child Abuse & Neglect, 104700–104700. PubMed. https://doi.org/10.1016/j.chiabu.2020.104700

